# Associations of individual exposure to particulate matter with next-day physical activity and sedentary behaviour in older adults: a micro-longitudinal study

**DOI:** 10.64898/2026.09.23.26363783

**Authors:** Mikaela Bloomberg, Maya Jones, Laura Brocklebank, Paola Zaninotto, Andrew Steptoe

## Abstract

**Background:** Staying physically active is essential for healthy ageing. Exposure to high levels of air pollutants such as particulate matter (PM) is associated with less active, more sedentary lifestyles in older adults. Whether results extend to typical fluctuations in daily PM exposure is unclear. We examined associations of higher-than-usual personal exposure to PM (PM_2.5_ and PM_10_) with next-day movement behaviour in older adults, assessed using concurrently worn personal air quality monitors and accelerometers.

**Methods:** The present study used 304 days of data from 67 adults aged 50-85 years living in the UK. For five days, participants wore accelerometers 24-hours per day and personal air quality monitors during waking hours. Average daily time spent in movement behaviours (moderate-to-vigorous physical activity [MVPA], light physical activity [LPA], sedentary behaviour [SB]) and step count was extracted from accelerometers, and average daily exposure to PM_2.5_ from personal air quality monitors. PM_10_ was examined in secondary exploratory analyses. We used linear mixed models to examine within-person associations of deviation from usual PM exposure with next-day movement outcomes, with estimates presented for ‘usual’ (individual mean) and ‘higher-than-usual’ (+1 interquartile range, equivalent to 2.6 μg/m^3^ beyond individual mean PM_2.5_ exposure and 2.8 μg/m^3^ beyond individual mean PM_10_ exposure) exposure. Analyses were repeated using area-based PM estimates.

**Results:** Compared with usual exposure, higher-than-usual PM_2.5_ was associated with 8 minutes less LPA (95% CI=-14 to –2) and 483 fewer steps (–855 to –82) on the following day. Exposure to higher than usual PM_10_ was associated with 8 minutes less LPA (–13 to –3), 8 minutes more SB (0 to 15), and 412 fewer steps (–739 to –85) on the following day. There was no association with MVPA, and no associations when area-level PM estimates were used as opposed to estimates from personal air quality monitors.

**Conclusions:** Typical daily variability in PM exposure in a setting with relatively low overall PM levels was associated with modest next-day differences in movement behaviour in older adults. Higher-than-usual PM exposure was associated with less next-day activity and more sedentary time. Personal monitoring may be important for capturing short-term exposure-behaviour associations.

**Trial registration:** N/A

## Background

Maintaining healthy movement behaviour patterns, including engaging in physical activity and limiting sedentary time, is essential for preserving physical function, independence, and wellbeing later in life. As movement becomes more variable with age (1), identifying environmental factors that may influence day-to-day activity patterns is increasingly important for supporting healthy ageing and preventing functional decline (2). Air pollution, and specifically particulate matter (PM), is an established determinant of health and mortality in older adults and has consistently been linked to lower levels of physical activity in large scale studies (3, 4). PM might also be related to shorter-term behavioural changes: acute PM exposure is associated with exacerbations of respiratory and cardiovascular conditions (5) as well as subclinical cardiovascular impairment and systemic inflammation (6), with some evidence of lagged effects (7). These physiological responses may unfold over hours and persist into the next day, contributing to symptoms such as fatigue, malaise, or discomfort (8) and potentially reducing motivation or physical capacity for everyday movement even among otherwise healthy individuals.

Despite plausible short-term pathways, it remains unclear whether typical day-to-day increases in PM—defined as deviations from an individual’s usual exposure—are associated with subsequent changes in movement behaviours. Limited evidence on short-term, day-to-day associations suggests a reduction in physical activity concurrent with or immediately following high PM exposure (9–13). However, these studies come from high pollution settings (12, 13) or clinical cohorts (9–11), limiting generalisability to older adults living in comparatively low ambient pollution settings by international standards, such as the UK. These studies also focus on absolute PM levels rather than within-person deviations from usual exposure. Finally, previous studies rely on area-level measures of PM, which do not capture variability in individual exposure arising from personal behaviour, indoor air pollutants, and local microenvironments (14).

To address these limitations, we examined associations between PM (PM_2.5_ and PM_10_) and next-day movement behaviour patterns using 304 days of personal PM monitoring and concurrent wrist-worn accelerometer data from 67 adults living in the UK aged 50 years and older. We examined whether deviations from usual personal PM exposure were associated with movement behaviours (moderate-to-vigorous physical activity [MVPA], light physical activity [LPA], sedentary behaviour [SB]) and step count on the following day.

## Methods

### Data sources

A convenience sample of 89 ambulant adults aged ≥50 years living in the UK and with internet access were recruited via Age UK networks, university mailing lists for older adults, and word of mouth through researchers’ personal networks to participate in a micro-longitudinal pilot study examining day-to-day relationships between personal PM exposure and movement behaviour. Data were collected between May and October of 2025.

Participants were sent a study pack in the post containing a wrist-worn accelerometer and wearable personal air quality monitor. Participants were asked to start wearing the devices on receipt (day 0). We then analysed the subsequent five full days (00:00-23:59, days 1-5) to ensure a consistent wear window across participants, with devices returned by post on day 6. Raw accelerometry and air pollution data files were clipped to match the start and end of the wear period prior to analysis. Participants were not given feedback on their activity or air pollution levels until after the data collection period finished.

### Movement behaviours

The Matrix 003 (Beijing XMatrix Tech. Co., Ltd, Beijing, China) is a wrist-worn triaxial accelerometer including a gyroscope and heart rate monitor that has previously been used to objectively measure movement behaviours in ∼20,000 participants in the China Health and Retirement Longitudinal Study (15). The Matrix 003 is comparable to the Axivity AX3 (16), which was worn by >100,000 participants in the UK Biobank (Doherty et al., 2017) and >20,000 participants in the China Kadoorie Biobank (Chen et al., 2023). Participants were asked to wear the device on their non-dominant wrist for 24 hours per day for five days and five nights. They were informed that they could wear the device in the shower, but it should be removed for prolonged water exposure (e.g., swimming) or extreme temperatures. The Matrix was set to have a sampling rate of 50 Hz with a dynamic range of ±8 gravitational units (*g*).

Raw acceleration data was processed using open-source algorithms developed and validated by the Oxford Wearables Group. Daily time spent in movement behaviours (MVPA, LPA, and SB) was produced using the *Biobank Accelerometer Analysis Tool* (https://github.com/OxWearables/biobankAccelerometerAnalysis, v7.1.1) (17). Daily step count was produced using *stepcount* (https://github.com/OxWearables/stepcount, v3.17.1) (18, 19). Data processing was done in line with largescale studies including the UK Biobank and China Kadoorie Biobank (20, 21). Participants were included if they contributed ≥3 days with valid data within each one-hour period of the 24-hour cycle. Non-wear was defined as uninterrupted periods of ≥60 minutes during which the standard deviation of acceleration on each axis was <13 m*g*. Participants were also excluded if the data could not be parsed, the device could not be calibrated, >1% of readings were ‘clipped’ (i.e., fell outside ±8 *g*) before or after calibration, or the average acceleration was implausibly high (>100 m*g*). Days with less than 22 hours of wear time were excluded and time spent in movement behaviours was rescaled to 24 hours for all participants.

### Air pollution

Daily personal PM exposure was measured using the Atmotube PRO (Atmotech Inc., San Francisco, USA), a small and lightweight light-scattering optical particle counter that estimates mass concentrations of particles with an aerodynamic diameter of <1 µm, <2.5 µm and <10 µm (PM_1_, PM_2.5_, and PM_10_, respectively). In an independent 14-week outdoor collocation study performed in accordance with US Environmental Protection Agency guidelines, Atmotube PRO units showed good correlation with a reference-grade monitor for hourly PM_2.5_ (R = 0.86) and moderate correlation for PM_10_ (R = 0.49) (22). Participants were instructed to wear the Atmotube PRO during waking hours, attaching it to their clothing using the provided clip or lanyard, and to place the device in their bedroom overnight. They were informed that the device was not waterproof. The Atmotube PRO was set to record PM every 15 minutes to ensure sufficient battery for the data collection period. Daily PM exposure was calculated as the mean of all 15-minute measurements across the 24-hour period, including overnight bedroom measurements. Overnight measurements were included because indoor air pollution during sleep is plausibly a substantial contributor to daily PM exposure.

For comparison with personal PM estimates, daily area level measures of PM_2.5_ and PM_10_ were drawn from the nearest Department for Environment, Food and Rural Affairs (DEFRA) (23) monitoring station to each participant’s residential postcode. To facilitate comparison with DEFRA, we report result for PM_2.5_ and PM_10_ in the present study. As low cost sensors have limited ability to distinguish PM_10_ from smaller particles (24), PM_10_ analyses were considered secondary and exploratory in nature.

### Covariates

Participants were asked to report demographic (age, sex), socioeconomic (education level), and health (self-rated health, chronic conditions) data via online survey at the start of the wear period. Participants were also asked whether they experienced mobility limitations (difficulty walking 100 yards, getting up from a chair, climbing a flight of stairs, stooping/kneeling/crouching, lifting/carrying 10 lbs, picking up a 5p coin, reaching/extending the arms, pulling/pushing large objects).

Characteristics included in the present analysis included age in years and gender (man, woman, or non-binary/other). We also included time-varying variables that were plausibly related to air pollution and activity patterns, including day of the week, temperature, and precipitation. Daily mean temperature and precipitation were extracted from the Weather Underground historical archive for each participant’s residential location, which aggregates observations from a large network of citizen weather stations (25).

### Statistical methods

We used linear mixed models to examine associations of deviation from usual PM exposure with next-day time spent in movement behaviours and step count, with separate models for each outcome (time spent in MVPA, LPA, SB, or step count) and particulate size (PM_2.5_ or PM_10_). Linear mixed models use all available data and accommodate missingness in the outcome under the missing-at-random assumption (26). These models included a random intercept at the individual level to account for intraindividual correlation of repeated observations; each participant contributed up to four observations. Because personal PM exposure is partly behaviourally generated (e.g., outdoor physical activity can increase measured exposure), same-day associations between PM and movement behaviours can reflect bidirectional coupling and are difficult to interpret causally. We therefore specified next-day movement behaviours as the primary outcomes and examined same-day PM-movement associations as a secondary analysis to assess the potential influence of behavioural coupling.

To determine whether fluctuations from usual exposure to air pollutants were associated with next-day movement behaviour, we examined within-person associations of air pollution on day *d* with movement behaviour and step count measured on day *d + 1*. At each time point, the exposure was calculated as 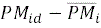, where *pM_id_* denotes the PM exposure of individual *i* on day *d*, and 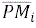 is the average PM exposure across the study period for individual *i*. We tested for non-linear associations by including quadratic and cubic terms for the exposure and retaining higher-order terms where significant (Wald test). Quadratic terms were retained in models for MVPA and step count.

Models also included a priori confounders as fixed effects. Models were adjusted for 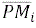 to partition between– and within-person effects. In this context, the within-person coefficient is not biased by time-invariant confounders, which are accounted for by adjustment for 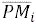. As such we included a minimal set of time-invariant covariates (age and sex) to improve precision of estimates only. Time-varying confounders included were those plausibly related to PM at *d* and movement behaviours at *d + 1*. They were drawn from day *d* and included day of the week as a categorical variable (which is deterministically related to the day of the week at *d + 1*) and weather (temperature and precipitation at *d*). Temperature and precipitation were included as these are related to air pollution at *d*, and activity at *d + 1* to the extent they reflect underlying weather trends. Mean temperature at *d* was fitted continuously using restricted cubic splines, and precipitation at *d* as a binary variable (yes or no) after examining including precipitation continuously and using splines.

To facilitate interpretation of the results, we present the continuous association as the estimated difference in next-day movement behaviour between an individual’s usual PM exposure (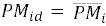) and ‘higher-than-usual’ exposure to PM. ‘Higher-than-usual’ exposure was defined as the participant’s usual exposure plus one sample-level IQR of the within-person deviation term, where the IQR was calculated on 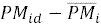 pooled across all participants and all person-days (i.e., not participant-specific). For linear models, reported estimates therefore correspond directly to the continuous model coefficient scaled to one sample-level IQR. For models including quadratic terms (MVPA and step count), reported estimates represent the difference in predicted outcome between a PM deviation of zero and one sample-level IQR, incorporating both the linear and quadratic terms. This scaling reflects a typical within-person increase above usual exposure. We also reported p-values for these estimates based on the Wald test, or a joint Wald test where quadratic terms were included (MVPA, step count). Analyses were performed in StataMP 18.0 or R version 4.4.2, with two-sided p-values<0.05 considered statistically significant.

### Additional analyses

Physical activity and sedentary behaviour are part of the 24-hour activity cycle along with sleep, where more time spent in one behaviour necessarily displaces time in another, and health impacts depend on the way in which the balance of behaviours shifts. To take this dependency into consideration, we first repeated analyses using compositional data analysis to determine whether deviations from usual personal PM exposure were associated with shifts in the next-day movement behaviour composition (comprising MVPA, LPA, SB, and sleep time on the following day, with sleep time also derived from raw acceleration data using the *Biobank Accelerometer Analysis Tool*). Details are provided in the Supplementary materials (Methods S1).

Second, to evaluate whether results differed between individual-level versus area-level exposure estimates, we repeated analyses using PM concentration assigned to each individual from the nearest DEFRA monitoring station. DEFRA measures outdoor ambient air, whereas the Atmotube PRO captures both indoor and outdoor personal exposure. DEFRA monitoring methods also incorporate measures to minimise relative humidity or moisture interference that are not used by the Atmotube PRO.

Finally, personal air pollution exposure on day *d* may be influenced by movement behaviour at the same time point, which could plausibly affect movement behaviour on day *d + 1.* We did not adjust for activity at *d* in the main analyses, as lagged outcomes are correlated with the individual-level random intercept. Their inclusion would therefore violate the random effects independence assumption, potentially biasing estimates. To determine whether same-day associations between PM exposure and movement behaviour influenced our results, we instead examined same-day associations with movement behaviour and step count.

## Results

Of 89 adults enrolled in the study, 1 participant withdrew and 1 did not return the devices. Of the remaining 87 participants, 67 (77.0%) had valid accelerometer and air pollution data, with 304 days of data in total (median follow-up, 5 days; IQR, 4-5). Missing data occurred due to random device malfunction (e.g., memory/storage issues or water damage).

Participant characteristics are presented in Table 1. The mean age in the analytic sample was 66 years (SD, 9.3) and 62.7% were women. Participants were highly educated (32.8% to university degree and 37.3% above university degree), mostly retired (65.7%), and in good health (47.8% reported very good or excellent health; no participants reported poor health); 31.3% nonetheless reported mobility limitations. Participants were primarily located in Greater London (86.6%). Each day, participants spent an average of 54 minutes (SD, 36 minutes) in MVPA, 3 hours and 54 minutes in LPA (SD, 1 hour 36 minutes), and 11 hours and 30 minutes in SB (SD, 1 hour 42 minutes). The median step count was 10,071 (IQR, 6405-14,097). Median exposure to PM_2.5_ during the study period was 5.0 μg/m^3^ (IQR, 3.7-7.8) and median exposure to PM_10_ was 6.8 μg/m^3^ (IQR, 5.3-10.0).

**Table 1.**
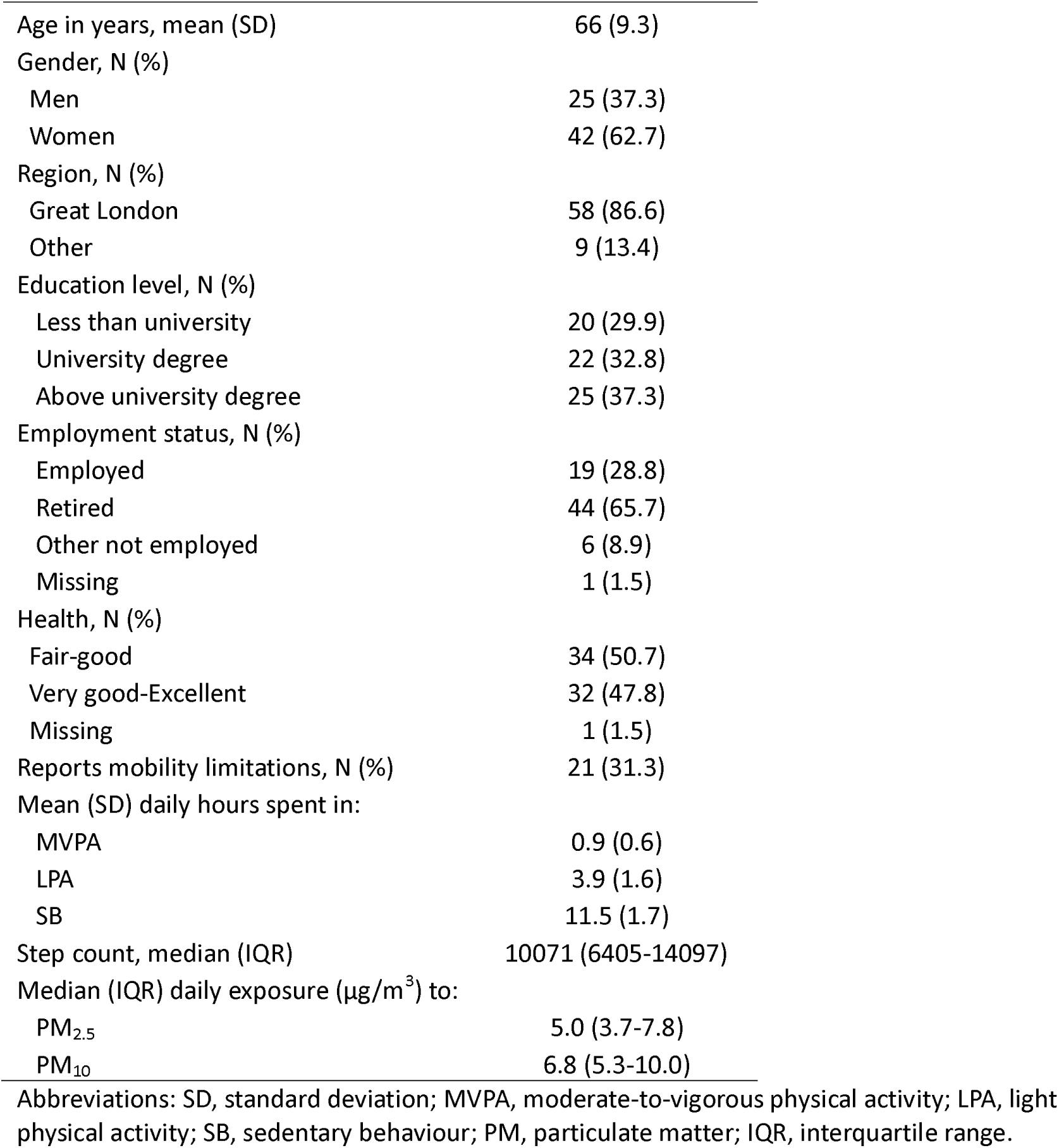
Participant characteristics (N=67).

In the following sections, we present results for ‘higher-than-usual’ compared with ‘usual’ PM exposure. In the analytic sample, ‘higher-than-usual’ PM exposure corresponded to 2.6 μg/m^3^ for PM_2.5_ and 2.8 μg/m^3^ for PM_10_. For example, for a participant with usual PM_2.5_ exposure of 5.0 μg/m^3^, this contrast compares predicted next-day movement behaviour at 5.0 and 7.6 μg/m^3^; for a participant with usual PM_10_ exposure of 5.0 μg/m^3^, this contrast compares 5.0 and 7.8 μg/m^3^. Separate linear and quadratic coefficients for models including quadratic terms are presented in the Supplementary materials (Table S1).

In general, higher-than-usual PM_2.5_ and PM_10_ exposure was associated with less LPA, more SB, and fewer steps on the following day (Table 2). Exposure to higher-than-usual PM_2.5_ was associated with 8 minutes less (95% CI, –14 to –2; p=0.01) LPA and 483 fewer steps (95% CI, –885 to –82; p=0.02) on the following day compared with usual exposure. Higher-than-usual PM_2.5_ also corresponded to 6 minutes more SB (95% CI, –2 to 15) on the following day, but the association did not reach statistical significance (p=0.15). The difference in next-day MVPA between usual and higher-than-usual PM_2.5_ exposure was comparatively minor (–3 minutes, 95% CI, –6 to 1) and did not reach statistical significance (p=0.20).

**Table 2.** Associations of deviation from usual particulate matter exposure with next-day movement behaviours and step count.

|  | **PM<sub>2.5</sub>** |  | **PM<sub>10</sub>** |  |
| --- | --- | --- | --- | --- |
|  | *Coefficient (95% CI)* | *P-value* | *Coefficient (95% CI)* | *P-value* |
| *MVPA* | -3 (-6 to 1) | 0.20 | -2 (-5 to 2) | 0.34 |
| *LPA* | -8 (-14 to -2) | 0.01 | -8 (-13 to -3) | 0.004 |
| *SB* | 6 (-2 to 15) | 0.15 | 8 (0 to 15) | 0.04 |
| *Step count* | -483 (-884 to -82) | 0.02 | -412 (-739 to -85) | 0.01 |

Movement behaviours are in minutes/day. Step count refers to number of steps/day. Unit for PM is per IQR change relative to usual exposure (IQR=2.6 µg/m<sup>3</sup> for PM<sub>2.5</sub>; 2.8 µg/m<sup>3</sup> for PM<sub>10</sub>). Based on linear mixed models adjusted for age, sex, day of the week, temperature, and precipitation.
Abbreviations: MVPA, moderate-to-vigorous physical activity; LPA, light physical activity; SB, sedentary behaviour; PM, particulate matter; CI, confidence interval; IQR, interquartile range.

Compared with usual exposure to PM_10_, exposure to higher-than-usual PM_10_ was associated with 8 minutes less (95% CI, –13 to –3; p=0.004) LPA on the following day. Higher-than-usual PM_10_ was also associated with 8 minutes more SB (95% CI, 0-15; p=0.04) and 412 fewer steps (95% CI, –739 to –85; p=0.01) on the following day. Similar to PM_2.5_, the difference in next-day MVPA between usual and higher-than-usual PM_2.5_ exposure was relatively small (–2 minutes, 95% CI, –5 to 2) and did not reach statistical significance (p=0.34).

Figure 1 shows estimated differences in next-day time use between individuals with usual and higher-than-usual PM exposure when accounting for the interdependence of 24-hour movement behaviours using compositional data analysis. Consistent with the main analyses, higher-than-usual PM exposure was associated with a shift in the overall next-day movement behaviour composition. This shift was characterised primarily by less LPA and more SB. For PM_2.5_, the estimated differences were 7 fewer minutes (–11 to –1) of LPA and 7 more minutes (–2 to 21) of SB the following day. For PM_10_, the estimated differences were 7 fewer minutes of LPA (–11 to –2) and 8 more minutes of SB (0 to 22). Estimated differences in MVPA and sleep time were minor (0-1 minutes).

**Figure 1.**
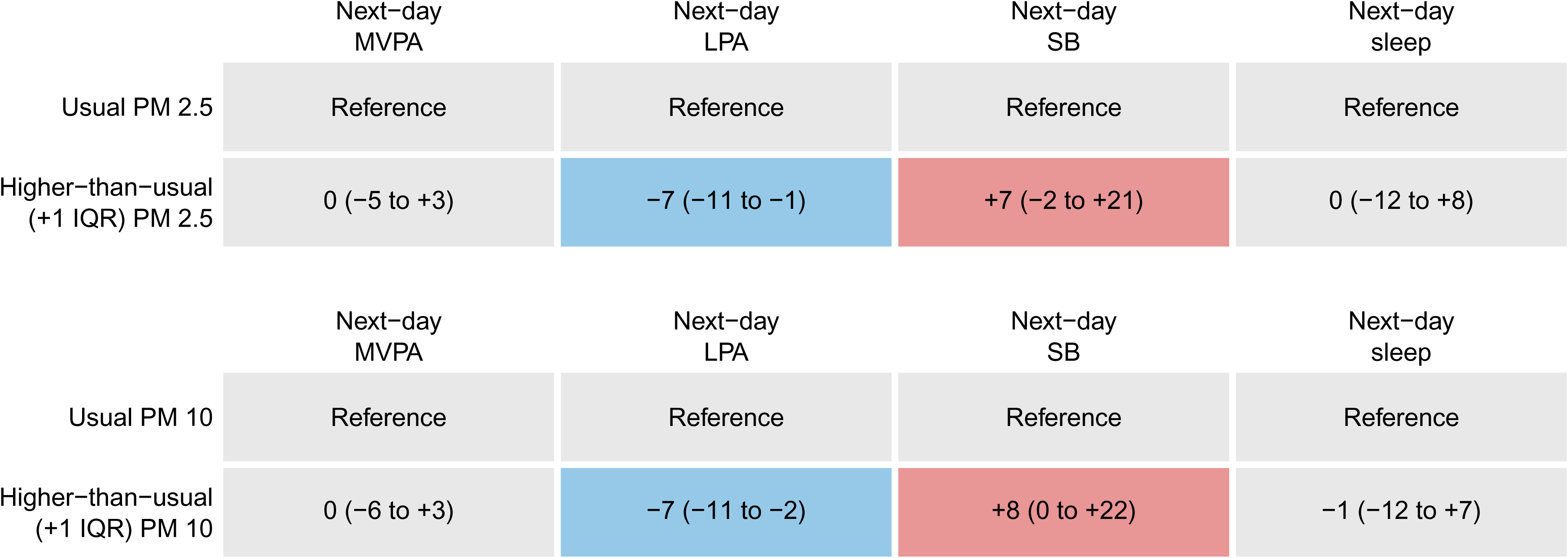
Estimated differences in next-day movement behaviours associated with higher-than-usual exposure to particulate matter. Predicted average differences (95% confidence interval) in minutes per day spent in each movement behaviour, relative to the reference value (individual mean exposure to given air pollutant during the study period). IQR for PM_2.5_ is 2.6 μg/m^3^; IQR for PM_10_ is 2.8 μg/m^3^. Positive values indicate more time in the given behaviour compared to the reference and negative values indicate less. Estimates are derived from compositional models adjusted for age, sex, day of the week, temperature, and precipitation. Abbreviations: MVPA, moderate-to-vigorous physical activity; LPA, light physical activity; SB, sedentary behaviour; IQR, interquartile range; PM, particulate matter.

When considering DEFRA area-level estimates rather than personal PM monitoring, median PM exposure over the study period was 5 μg/m^3^ (IQR, 4-7) for PM_2.5_ and 10 μg/m^3^ (IQR, 8-13) for PM_10_. Correlation between DEFRA and Atmotube PRO estimates was 0.27 for PM_2.5_ and 0.21 for PM_10_. In contrast to the main analyses, within-person deviations from usual DEFRA-estimated PM exposure were not related to next-day movement behaviours or step count (Table S2). For example, higher-than-usual PM corresponded to 2 minutes more (–6 to 9) LPA on the following day for PM_2.5_ (p=0.64) or 3 minutes more (–7 to 14) LPA for PM_10_ (p=0.51). Finally, deviation from usual pollutant exposure was not related to same-day movement behaviours or step count (Table S3).

## Discussion

In this micro-longitudinal study of daily air pollution exposure and next-day movement behaviours undertaken using 304 days of data from 67 UK-based adults aged 50-85 years, higher-than-usual exposure to PM was associated with less physical activity and more sedentary behaviour on the following day. DEFRA pollution estimates did not show the same associations with next-day activity patterns. Taken together, these findings suggest that typical daily variation in personal PM exposure may be associated with next-day movement patterns, and that this signal may be missed when exposure is assigned using area-level monitoring data.

Previous work linking higher daily air pollution exposure to reductions in physical activity has focused on same-day associations and relied on area-level exposure assignment (9, 10, 12, 13), and in one case, subjective report of physical activity (12). Evidence on lagged effects is more limited, but a study in 408 older adults with chronic obstructive pulmonary disease reported reduced physical activity several days after high pollution exposure (11). Extending this literature, we examined within-person deviations from usual personal PM exposure in older adults without chronic respiratory disease and found that typical day-to-day increases above an individual’s usual exposure were associated with next-day shifts in physical activity and sedentary behaviour and in the 24-hour movement behaviour composition. Notably, when we reassigned exposure using area-level DEFRA estimates, we did not observe the same next-day associations, suggesting that ambient assignment may not capture behaviourally relevant variation in personal exposure. The relatively low correlations between DEFRA and Atmotube PRO estimates further highlight that area-level ambient and personal monitoring capture different aspects of PM exposure. Associations were observed for LPA and SB, whereas MVPA was largely unchanged. This pattern is consistent with evidence that suggests people do not change their physical activity routines in response to air pollution until it reaches very high levels (27). PM levels in this study were low in absolute terms, which may have been insufficient to deter planned activity but sufficient to shift day-to-day discretionary movement.

One possible explanation for the observed next-day changes in movement behaviour is a delayed subclinical physiological response to PM exposure. Short-term PM_2.5_ exposure has been associated with systemic inflammation, oxidative stress, and transient reductions in lung function, although much of this mechanistic evidence comes from settings with substantially higher PM_2.5_ concentrations than those observed in the present study (6, 28). At the same time, relatively low PM_2.5_ concentrations should not be interpreted as biologically irrelevant: although concentrations in our study were generally below the WHO 24-hour air quality guideline, this guideline does not represent a threshold below which adverse effects are absent, and short-term exposure in settings with relatively low ambient concentrations has been associated with adverse clinical outcomes (29). Whether fluctuations in PM_2.5_ within relatively low ambient PM settings can also produce subclinical physiological responses that are sufficient to influence subsequent movement behaviour is not yet established and warrants further investigation.

There are also relevant non-causal explanations for the findings. It is possible that because individual exposure to pollutants is partially behaviourally driven, more activity on one day could drive higher-than-usual same-day exposure and correspond to less activity on the following day. Our analyses did not show clear same-day associations between movement behaviours and PM deviations, suggesting that this is less likely to be the sole driver of the next-day pattern. However, because exposure and activity may influence each other in opposite directions within the same day, a null same-day association does not entirely eliminate the possibility that behaviourally driven exposure contributes to the observed lagged association. Other unmeasured day-to-day routines or planned activities could also jointly shape personal PM exposure and subsequent movement behaviour; longer studies collecting more information on daily context are needed to distinguish between these explanations and strengthen causal inference.

The main strength of our study is the micro-longitudinal study design and use of personal air quality monitoring to capture individuals’ personal pollutant exposure as they moved through their daily routines, rather than relying on area-level pollution estimates. Personal monitoring also captures substantial between-person variability driven by indoor microenvironments and time-activity patterns—sources of exposure that are largely missed by area-level ambient estimates. This personal monitoring was combined with wrist-worn accelerometry to provide continuous, objective measurement of movement behaviours throughout the 24-hour day, without relying on self-report. Finally, although the sample size was modest, repeated observations over five days per participant enabled examination of within-person day-to-day associations between deviations in exposure and next-day movement behaviours.

There are several limitations to this study. Our sample was a convenience sample of adults aged 50 years and older who volunteered to wear both devices for five days, where participants were highly educated, in good self-reported health, and relatively active. It is possible that different associations would have been observed with a larger or more heterogeneous sample and replication is needed. While time-invariant confounding is accounted for by adjustment for individual mean exposure, residual time-varying confounding could nonetheless affect causal inference. Due to battery limitations, the Atmotube PRO personal air quality monitor was set to measure PM concentrations at 15-minute intervals rather than continuously, potentially missing brief exposure peaks. In line with the lower correlation observed for PM_10_ than for PM_2.5_ in collocation testing, PM_10_ estimates may be noisier or less accurate. More broadly, low-cost optical sensors have limited ability to distinguish particles in the coarse fraction (24), meaning that PM_10_ estimates may largely reflect the smaller-particle contribution to PM_10_ and should not be interpreted as an independent measure of coarse particulate exposure. Although each participant used the same sensor throughout the monitoring period, device-specific differences in sensitivity and within-device measurement error could affect the magnitude and precision of estimated within-person changes in PM exposure, and consequently the estimated associations. Atmotube PM estimates can be affected by very high (e.g., >80%) relative humidity (22); however, humidity levels in this study were well below the range where performance is reported to degrade (99th percentile 64%), making substantial humidity-related bias unlikely.

Ambient exposure assignment in this study was based on monitoring-station estimates at residential location and may not reflect more refined modelled ambient exposures. However, even high-resolution ambient models tied to residential addresses may not fully capture behaviour-driven variation in personal exposure. Finally, the short monitoring period (up to five days per participant) limited our ability to examine multi-day lag structures (e.g., distributed lag models) or to fit more complex dynamic within-person models; longer monitoring is needed to characterise the timing of effects more fully.

## Conclusions

The findings of the present study suggest that typical day-to-day increases in PM exposure relative to an individual’s usual levels are associated with measurable reductions in LPA. The 8-minute difference in LPA corresponds to approximately 3% of mean daily LPA in the sample, while the observed difference in step count corresponds to approximately 4-5% of the median daily step count. While the observed fluctuations were relatively modest, even minor daily shifts in light activity may be meaningful in older adults, as modest reductions in activity can accumulate over time and contribute to greater sedentary exposure. While MVPA is often the main focus of public health recommendations, meta-analyses suggest that LPA is also strongly associated with risk of mortality in middle-aged and older adults (30). LPA also represents the largest proportion of daily movement in older adults, as MVPA decreases and LPA increases with age (31). Our findings therefore highlight an important area for future research to understand how typical variations in air pollution might influence routine activity in older adults, beyond extreme pollution events. Finally, these results demonstrate the potential of personal air quality monitors to capture individual air pollutant exposure patterns, whereas area-level measures may miss behaviourally relevant variation in exposure. However, given the sample size and short monitoring period, replication in larger, more diverse samples with extended monitoring is warranted.

## Declarations

### Ethics approval and consent to participate

The study was approved by the University College London Research Ethics Committee (Reference: 28517/001). Participants provided written informed consent. All data collection was performed in accordance with the Declaration of Helsinki.

### Consent for publication

Not applicable.

### Availability of data and materials

The dataset generated and analysed during the current study are not publicly available due to institutional data sharing restrictions but are available from the corresponding author on reasonable request.

### Authors’ contributions

Conceptualisation: MB, PZ, AS.

Data curation: MB, MJ.

Formal analysis: MB.

Funding acquisition: MB, PZ, AS.

Investigation: MB, MJ.

Methodology: MB, LB, PZ.

Project administration: MB, MJ.

Supervision: PZ, AS.

Visualisation: MB

Writing – original draft: MB, MJ.

Writing – review and editing: All authors.

## Abbreviations

MVPA: Moderate-to-vigorous physical activity
LPA: Light physical activity
SB: Sedentary behaviour
IQR: Interquartile range
CI: Confidence interval
PM: Particulate matter
DEFRA: Department for Environment, Food & Rural Affairs
CoDA: Compositional analysis of data
ILR: Isometric log-ratio

## Acknowledgements

Not applicable.

