## Supplementary materials for "Associations of individual exposure to particulate matter with next-day physical activity and sedentary behaviour in older adults: a micro-longitudinal study"

**Supplementary methods**

Methods S1. Compositional data analysis.

We applied compositional analysis of data (CoDA) to determine whether deviations from usual personal PM exposure were associated with shifts in the 24-hour movement behaviour composition (comprising moderate-to-vigorous physical activity [MVPA], light physical activity [LPA], sedentary behaviour [SB], and sleep time) on the following day.

CoDA is a well-established method for analysing compositional data, where the parts sum to a whole (e.g., 24 hours in a day) (1-3). It converts the proportions of time spent in each behaviour into isometric log-ratio (ILR) coordinates, which express the relative balance of one behaviour compared to others in a mathematically appropriate way for compositional data. This transformation avoids statistical issues that arise when analysing raw proportions directly due to their inherent interdependence. While ILRs themselves are not interpretable, they allow us to perform appropriate regression analysis on compositional data, after which results can be translated back into meaningful time-use differences. As compositional data cannot include zeros, we used multiplicative replacement to impute small values (3.6 seconds) for zeros, as has been done previously (4).

We used the *ilr* function from the *compositions* package (2) in R to convert time-use data into ILR coordinates. This function constructs ILRs by sequentially comparing one behaviour to the remaining set. We defined three ILRs: 1) MVPA relative to LPA, SB, and sleep; 2) LPA relative to SB and sleep; and 3) SB relative to sleep. Each ILR coordinate was then used as the dependent variable in a separate linear mixed effects model, with $PM_{id} - \overline{PM}_i$ as the predictor and a random intercept at the individual level. Here, $PM_{id}$ denotes the PM exposure of individual $i$ on day $d$, and $\overline{PM}_i$ is the average PM exposure across the study period for individual $i$.

Because ILRs are not directly interpretable, we illustrated model results by back-transforming predicted ILR values into estimates of time spent in each movement behaviour using standard procedures (2). These predictions were generated for PM values corresponding to 'usual' exposure to PM ($PM_{id} = \overline{PM}_i$) and 'higher-than-usual' exposure to PM. 'Higher-than-usual' exposure was defined as one IQR higher than the participant's usual, where the IQR refers to the distribution of day-to-day deviations from participants' own means ($PM_{id} - \overline{PM}_i$) pooled across all person-days. This scaling reflects a typical within-person increase above usual exposure. We also calculated the model-predicted differences in time use between 'usual' and 'higher-than-usual' PM exposure. Confidence intervals around the predicted time use estimates were derived via nonparametric bootstrapping with 1000 replications; such confidence intervals can be used to indicate precision of estimates but are not related to significance testing.

**Supplementary tables**

Table S1. Linear and quadratic coefficients from moderate-to-vigorous physical activity and step count models.

| | $PM_{2.5}$ | | $PM_{10}$ | |
|---|---|---|---|---|
| | *Coefficient (95% CI)* | *P-value* | *Coefficient (95% CI)* | *P-value* |
| *MVPA* | -4 (-8 to 1) | 0.11 | -2 (-6 to 1) | 0.22 |
| *$MVPA^2$* | 1 (0 to 2) | 0.006 | 1 (0 to 1) | 0.03 |
| *Step count* | -580 (-1032 to -129) | 0.01 | -475 (-836 to -114) | 0.01 |
| *Step $count^2$* | 97 (23 to 172) | 0.01 | 63 (6 to 119) | 0.03 |

MVPA unit is minutes/day. Step count unit is steps/day. Coefficients are presented on the sample-level interquartile range (IQR) scale used in the main analyses. Linear terms therefore represent a one-IQR increase in within-person PM deviation, and quadratic terms are scaled to $IQR^2$. Abbreviations: CI, confidence interval; MVPA, moderate-to-vigorous physical activity.

Table S2. Associations of deviation from usual particulate matter exposure (based on DEFRA estimates) with next-day movement behaviours and step count.

| | **$PM_{2.5}$** | | **$PM_{10}$** | |
|---|---|---|---|---|
| | *Coefficient (95% CI)* | *P-value* | *Coefficient (95% CI)* | *P-value* |
| *MVPA* | -4 (-8 to 0) | 0.07 | -2 (-7 to 3) | 0.44 |
| *LPA* | 2 (-6 to 9) | 0.64 | 3 (-7 to 14) | 0.51 |
| *SB* | 2 (-2 to 5) | 0.76 | -5 (-19 to 10) | 0.51 |
| *Step count* | -283 (-701 to 133) | 0.18 | -111 (-667 to 444) | 0.70 |

Movement behaviours are in minutes/day. Step count refers to number of steps/day. Unit for PM is per IQR change relative to usual exposure (3.0 µg/m$^3$ for $PM_{2.5}$; 5.0 µg/m$^3$ for $PM_{10}$). Air pollutant concentrations are drawn from DEFRA estimates based on participant residential post code. Based on linear mixed models adjusted for age, sex, day of the week, temperature, and precipitation. Abbreviations: MVPA, moderate-to-vigorous physical activity; LPA, light physical activity; SB, sedentary behaviour; IQR, interquartile range; PM, particulate matter; CI, confidence interval

Table S3. Associations of deviation from usual particulate matter exposure with same-day movement behaviours and step count.

| | $PM_{2.5}$ | | $PM_{10}$ | |
|---|---|---|---|---|
| | *Coefficient (95% CI)* | *P-value* | *Coefficient (95% CI)* | *P-value* |
| *MVPA* | -2 (-6 to 2) | 0.28 | -2 (-4 to 1) | 0.32 |
| *LPA* | 4 (-2 to 10) | 0.20 | 5 (0 to 10) | 0.05 |
| *SB* | 1 (-7 to 9) | 0.81 | -1 (-8 to 6) | 0.75 |
| *Step count* | 57 (-308 to 421) | 0.76 | 106 (-191 to 404) | 0.48 |

Movement behaviours are in minutes/day. Step count refers to number of steps/day. Unit for PM is per IQR change relative to usual exposure (2.6 $\mu g/m^3$ for $PM_{2.5}$; 2.8 $\mu g/m^3$ for $PM_{10}$). Based on linear mixed models adjusted for age, sex, day of the week, temperature, and precipitation.
Abbreviations: MVPA, moderate-to-vigorous physical activity; LPA, light physical activity; SB, sedentary behaviour; IQR, interquartile range; PM, particulate matter; CI, confidence interval
